# Serosurveillance of SARS CoV 2 among the healthcare workers of a tertiary care teaching institution in Central Kerala during the post lockdown phase

**DOI:** 10.1101/2021.01.27.21250502

**Authors:** Aboobacker Mohamed Rafi, Maglin Monica Lisa Joseph Tomy, Ronnie Thomas, Chithra Valsan, U G Unnikrishnan, Susheela J Innah, Praveenlal Kuttichira

## Abstract

**Background:** Kerala was the first state to have the confirmed case of COVID-19 in the country and it was first confirmed in Thrissur district on 30 January2020.Our institute being in the heart of the city had to take adequate measures to mitigate the spread and treat the required patients by keeping its staff safe & Healthy. The hallmark of COVID 19 infection is high infectivity, pre-symptomatic transmission and asymptomatic prevalence which could result in high cumulative numbers of infections, hospitalizations, and deaths. Kerala was the first state to confirm community transmission in July 2020.Health care workers being in the forefront in the war against COVID19 are very prone in acquiring the infection and are possible to be asymptomatic sources for cluster formation. Knowing the development of immunity as shown by the presence of anti COV2 antibodies in the population contributes to the epidemiological understanding of the disease. The intent of the study is to do an antibody testing in our hospital to find the serosurveillance of SARS CoV 2 among the healthcare workers in our hospital.

**Aim:** To estimate the seropositivity of SARS CoV 2 among the healthcare workers at Jubilee Mission Medical College and Research Institute, Thrissur, Kerala, six months after revoking the lockdown

**Methodology:** A cross sectional study among the health care workers of the medical college. Multistage Sampling was done with the hospital block as the first stage and departments as the second stage. In the final stage of sampling the test individuals were selected on a first come first served basis after the antibody test availability was declared open and free for all staff. A consent form and a Google form were given to all staff who volunteered for participating in the study. Each consented participant recruited into the investigation completed a questionnaire which covers details regarding demographics, exposure history, Residence & travel. Blood sample was collected and Anti-SARS COV2 IgG antibody testing which targets the Spike Protein 1(SP1) was done using the VITROS chemiluminescence platform (Orthoclinical diagnostics, USA). Sampling & testing ranged over a time frame from September 5^th^ to December 15^th^, 2020

**Results:** Jubilee Mission Medical College has 2785 working staff at the time of study. A total of 420 staff consented and their samples were tested. 37 staff members tested positive for COVID-19 antibody, yielding an overall prevalence of 8.75% (95% CI, 6.23–11.86). 86.5 % (32/37) of them were having a history of COVID-19 Antigen / RT PCR Positivity. We identified a statistically significant linear trend (p value =0.00001), between seropositivity and the degree of severity of COVID 19. Among the various factors which increase the risk of seroconversion, history of undergoing quarantine (p value < 0.001), contact with a confirmed case (p value = 0.002), contact with a caregiver for COVID 19 (p value =0.001) and history of Upper respiratory symptoms (p value =0.001), were found to be significantly associated with positive serology.

**Conclusions:** The overall seropositivity in the current study was found to be 8.75% which is comparable to seroprevalence studies conducted in the United States and Wuhan in China. The pattern of seropositivity across the different category of health workers observed in the present study showed a higher prevalence among nurses. This result is also in agreement with a recent published report from united states. Various measures advised by the national and state health authorities were adequately adhered to. Keeping track of the pattern of development of immunity in the community is part of understanding the illness and forecasting the spread. For the tested HCW, it will boost up morale by ending uncertainty. For the hospital administration it will help in decision making about relative focusing of interventions on patients in general and HCWs. By knowing the immunity status of HCWs, the Institution will be able to contribute authentically to the development of intervention strategies and guidelines from time to time, besides following the available guidelines. Being an educational institution, it is obligatory to train all the elements of care delivery to the future generation of health care workers. Getting experienced from a small but relevant sample was expected to facilitate larger community study envisaged in peripheral areas Jubilee served

## Introduction

The hallmark of SARS-CoV-2 pandemic is high infectivity, pre-symptomatic transmission and asymptomatic prevalence which results in high cumulative numbers of infections, hospitalizations and deaths.^1^ In India, Kerala was the first state affected by COVID-19, and the first coronavirus case was confirmed in Thrissur district on 30 January2020. ^2^ By early March, the state had the highest number of active cases in India mainly due to a huge number of cases imported from other countries and states. Using the five components of trace, quarantine, test, isolate and treat, by 10 June 2020, Kerala managed to keep the basic reproduction number at 0.454 against the India and world averages from 1.225 and 34.^3^

The state of Kerala has witnessed a long lag phase after the first reported case of COVID 19 on Jan 30 ^4^ till the phase of community spread and has managed to delay the peak of the pandemic by successful implementation of various control measures. In each phase starting from the reporting of the first case in India, our hospital along with other hospitals in the state have taken appropriate control measures to fight against the pandemic ^5^. It is important to have knowledge about the effectiveness of the steps taken, in planning the future strategies for disease control. Silent infection is a major concern. It is also important to know if health care workers act as a source of infection to patients during a pandemic, especially when hospitals serve other patients also. Being a novel virus, little is known about various aspects of the virus; the factors contributing to its spread, the progression of the disease, development of immunity etc. Knowing the development of immunity as shown by the presence of antibodies in the population contributes to the epidemiological understanding of the disease. Studies on seroconversion rates in a population helps to find out the exposure to the virus in that population, be it symptomatic or asymptomatic^6^. WHO continues to review the evidence on antibody responses to SARS-CoV-2 infection. Most of these studies show that people who have recovered from infection have antibodies to the virus. Seroprevalence among various categories of tested individuals reported from various parts of India ranged from 0.73 to 19.8^7-13^. So far, little is known about it related to Kerala. Seroprevalence studies conducted by the ICMR during the first two phases shows a gradual rise from 0.33 % in May to 0.8% in August 2020.^14^ Hence, we planned to undertake an antibody testing among health workers in our hospital.

### Aim

To estimate the seropositivity of SARS CoV 2 among the healthcare workers at Jubilee Mission Medical College and Research Institute, Thrissur, Kerala, six months after revoking the lockdown

### Objectives

We intended to find out the seropositivity in health care workers for COVID 19 during the rising graph of pandemic incidence in this part of Kerala, and to compare with reports from elsewhere in the country.

### Methodology

The Institutional Review Board approval was obtained for this study. The study was conducted among the HCWs at Jubilee Mission Medical College & Research institute, Thrissur, a 1600 bedded teaching hospital with around 3000 staff in the regular pay rolls, daily wagers and workers of service contractors. This study was designed as a cross sectional study. All categories of staff working in the hospital and medical college comprised the study population. A rising graph of incidence was operationally defined as a period up to daily incidence in the state above 10000 and or daily COVID In Patient strength in our hospital as 100 or above.

### Sample Size

Sample size was calculated by the formula n = (Zα) 2 ×p×q /d^2. Zα is the Z value at an α error of 0.05. i.e., 1.96 for a 95% confidence interval p, 23%, is the proportion of subjects with positive SARS COV2 antibodies according to a study done by Percivalle E et al during the peak of epidemic in Italy ^6^. q is 100-p. d is clinically allowable error which was taken as 20% of prevalence. The minimum sample size required was calculated to be 320

### Sampling Technique

Multistage Sampling was done with the hospital block as the first stage and departments as the second stage. In the final stage of sampling the test individuals were selected on a first come first served basis after the antibody test availability was declared open and free for all staff. A consent form and a Google form were given to all staff who volunteered for participating in the study. Research staff helped the volunteers to detail the consent form and filling up of the proforma in Google form. Each consented participant recruited into the investigation completed a questionnaire which covers demographic information, exposure history, Residence details (containment zone or not), travel and details of family exposure.

### Sample collection

After obtaining informed consent 3 ml of blood was collected in EDTA vacutainers, centrifuged and the plasma was separated. The plasma was subjected to antibody testing by Chemiluminescence Immunoassay. The kit used was manufactured by Ortho clinical diagnostics (USA). The kit was used in the Vitros equipment. The blood samples were tested for the presence of Anti SARS COV2 IgG antibody. Testing Time period: The process of Recruitment & Sampling started on 5th September 2020 and ended on 15th December 2020.

### Testing Methodologies

We used the chemiluminescence technology for antibody testing. These tests can target the Spike-protein S1 antigen, Spike-protein S2 antigen, nucleocapsid antigen, or a combination. The assay which we used in this study was Vitros anti-SARS-COV-2 IgG, which targets the S1 spike protein. As compared to other coronaviruses, S1 protein is more specific and unique to COVID-19. The test kit used in the present study has a sensitivity of more than 90% and specificity of nearly 100%. Lin et al reported the superiority of chemiluminescence-immunoassay over the ELISA method.^15^

The details and results of the tests done were recorded. For those HCWs who became positive in antibody testing, the details were shared with the institutional medical board and further necessary action if found was offered as per Kerala government guidelines and Institutional policy. Antibody positive status among tested samples were calculated and considered as seropositivity. It was calculated overall for all HCWs and separately for different categories of them.

### Statistical Analysis

For the summary of demographic characteristics, continuous variables were summarized as mean values and standard deviation while categorical variables were summarized as proportions. The χ2 test was used for comparing the epidemiological features between positive and negative cases. Chi square for trend analysis was done for exploring the relationship of the degree of severity with test positivity. All analyses were conducted using SPSS version 25.

## Observations and Results

Jubilee Mission Medical College & Research Institute has 2785 working staff at the time of study. A total of 423 staff (Table 1) consented and their samples were tested. This included 77 Doctors, 104 nursing staff, 85 technicians, 31 administrative /ministerial staff and 126 support staff. During the study period, 37 staff members tested positive for COVID-19 antibody, yielding an overall prevalence of 8.75% (95% CI, 6.23– 11.86). 86.5 % (32/37) of them were having a history of COVID-19 Antigen / RT PCR Positivity.

**Table1:** Test and result status of antibody to SARS-COV2 among Health Care Workers

| Category | Working | Tested | Positive | Seropositivity |
| --- | --- | --- | --- | --- |
| Doctors | 486 | 77 | 5 | 6.5 |
| Nurses | 978 | 104 | 12 | 11.5 |
| Technicians | 154 | 85 | 6 | 7.1 |
| Administrative Ministerial | 48 | 31 | 0 | 0.0 |
| Support | 1119 | 126 | 14 | 11.11 |
| **All** | **2785** | **423** | **37** | **8.75** |

The mean age of positive and negative employees was 35.35 and 34.46 years, respectively (Table 2). Employees who were females comprised a greater proportion of study subjects and there was no statistically significant difference in the seroprevalence (*P* = 0.327).

**Table 2:** Age & Gender

| Attribute |  | SARS CoV 2 Antibody |  | P value |
| --- | --- | --- | --- | --- |
|  |  | Positive (n=37) | Negative (n=386) |  |
| Mean Age |  | 35.35 ± 9.69 | 34.46 ± 10.59 | 0.624 |
| Gender | Male | 8 (29.3%) | 120 (21.6%) | 0.327 |
|  | Female | 29 (70.7%) | 266 (78.4%) |  |
| Table 2: Age & Gender |  |  |  |  |

The various risks for seroconversion were analyzed. Among the various factors which increase the risk of seroconversion, history of undergoing quarantine (p value < 0.001), contact with a confirmed case (p value = 0.002), contact with a caregiver for COVID 19 (p value =0.001) and history of Upper respiratory symptoms of COVID 19 (p value =0.001), were found to be significantly associated with positive serology.(Table 3)

**Table 3:** Chance of exposure and SARS COV 2 antibody status

| Sl.No | Attribute |  | SARS COV 2 Antibody |  | P value |
| --- | --- | --- | --- | --- | --- |
|  |  |  | Positive (n=37) | Negative (n=386) |  |
| 1 | History of symptoms suggestive of COVID 19 in the past 3 months | Yes | 26 | 91 | 0.001* |
|  |  | No | 11 | 295 |  |
| 2 | History of contact with a confirmed case/Quarantined individual | Yes | 24 | 147 | 0.002* |
|  |  | No | 13 | 239 |  |
| 3 | History of Travel outside Kerala since COVID outbreak | Yes | 1 | 9 | 0.997 |
|  |  | No | 36 | 377 |  |
| 4 | History of undergoing Quarantine | Yes | 27 | 85 | <0.001* |
|  |  | No | 10 | 301 |  |
| 5 | History of taking Covid 19 testing (RT-PCR/Antigen) | Yes | 30 | 160 | <0.001* |
|  |  | No | 7 | 226 |  |
| 6 | Adherence to PPE protocols of HICC | Yes | 35 | 370 | 0.715 |
|  |  | No | 2 | 16 |  |
| 7 | History of contact with a caregiver for Covid 19 patients | Yes | 24 | 142 | 0.001* |
|  |  | No | 13 | 244 |  |

## The degree of severity of symptoms and the antibody response was also analyzed

We identified a statistically significant linear trend (p value =0.00001), between seropositivity and the degree of severity of symptoms of COVID 19 (Table 4).

**Table 4:** Severity of symptoms and SARS COV 2 antibody status Chi-Square for trend. P value =0.00001

| Severity of Symptoms in past 3 months | SARS COV 2 Antibody Status |  | Odds ratio |
| --- | --- | --- | --- |
|  | Positive (n=37) | Negative (n=386) |  |
| Asymptomatic | 9 (24.3%) | 287 (76.1%) | 1 |
| Mild Upper Respiratory Symptoms | 8 (21.6%) | 69 (18.3%) | 3.22 |
| Moderate Symptoms | 17 (45.9%) | 29 (7.7%) | 18.62 |
| Severe | 3 (8.1%) | 1 (0.26%) | 95.33 |

## Discussion

The initial ‘Kerala model’ of response to the COVID-19 pandemic was to trace contact and quarantine or test, isolate and treat all travelers from foreign countries as well as neighboring states, coupled with lockdown and break the chain measures. This practice had made the state succeed in preventing community spread in the initial months of the Pandemic. With unlocking according to national policy and easing of restrictions, the community transmission was inevitable. Kerala was the first state in the country to declare community transmission in the Trivandrum district among the coastal region ^16^. There is always a high risk of transmission to the healthcare worker (HCW) from pre-symptomatic and asymptomatic patients reporting with non-COVID illness, especially in non-COVID-19 hospitals. Identifying infected HCWs, including asymptomatic ones, is important to reduce nosocomial spread^17^.

Initially a stratified random sampling was decided as the sampling technique. Risk groups were considered as the strata of the study. Hospital employees were stratified into four risk groups namely High risk, Moderate risk, Low risk and Very low risk based on the likelihood of exposure to a suspected COVID 19 patient, frequency and duration of exposure and the nature of interventions being performed. WINPEPI was used to select the target HCW. It was planned to study HCWs during the rising curve, plateau, falling, and post fall steady state of incidence graph. However, ‘no direct benefit to the tested individual’ was raised and honoring the voice of dissent, it was decided to withhold target individual selection and make it an open invitation to all HCWs for volunteering. The sample gathered were 423, one hundred in excess of initial target. This reflects the enthusiasm of the HCW in volunteering for a study which can have only collective benefit.

The overall seropositivity in the current study was found to be 8.75%. This estimate is comparable to that of seroprevalence studies conducted among general population in the United States and Wuhan in China which have reported a seroprevalence of 6.9% and 3.8% respectively ^18,19^. The prevalence of antibodies largely depends on the stage of the epidemic in the area at the time of the study. The findings of the present study contrast with the results of the population-based studies done in India. The nationwide study done in India in the early phase of the pandemic by the ICMR revealed a seroprevalence of 0.73% while the Sero-prevalence study across Delhi showed the prevalence of IgG antibodies to be 23.48 %.^20,21^ Studies done in similar settings among healthcare workers in Italy and India reported a seroprevalence of 14.4% and 11.94%. respectively ^22^. The variability in prevalence in a large country with multiple local communities having varying health status, demographic profile and ecology is understandable. The containment strategies adopted by the local Governments had to follow the National guidelines, but the efficiency of it varied. This also would explain the variation.

The pattern of seropositivity across the different category of health workers observed in the present study showed a higher prevalence among nurses. This result agrees with a recent published report from united states.^21^ These findings imply a higher occupational risk for SARS-CoV-2 infection among nurses. The Seropositivity was lowest among those working in nonclinical environments without patient contact. The history of upper respiratory symptoms in the past 3 months and a history of undergoing quarantine were significantly associated with a higher probability of SARS-CoV-2 specific antibody positivity. This result was found to be consistent with a similar report from a hospital in North India.^*23,24*^

We found a significant linear association with covid 19 antibody positivity and severity of disease with the likelihood of being seropositive being highest for those with severe respiratory infections. During the Initial phase of the pandemic, strategies that aimed to increase herd immunity by exposing young low-risk individuals to the virus were under consideration.^25^ But early evidence suggested that acquired immunity may be short-lived in individuals with mild or asymptomatic infections^26^. Further studies confirmed that a stronger antibody response was associated with disease severity^27^.

Sero-positivity among the staff of our Institution showed a comparatively low figure. Even after a period of 10 months after the first case detection in the district and 6 months after first case detection in our own hospital, the development of immunity against Covid-19 remaining low is a matter of concern. Only when herd immunity develops or enough people are vaccinated, the pandemic of viral disease will get controlled and eradicated.

Right from the beginning of this pandemic and since the reporting of the first COVID 19 case of India from Thrissur our hospital has adopted strict infection control measures. Various measures were adopted by our institute to contain the spread. The classroom teaching for MBBS and Nursing were suspended and shift to online mode ensured, though with certain concerns ^28,29^. A Multi-disciplinary Institutional Medical Board was constituted to take necessary actions regarding COVID positive patients and infection control among staff. The Hospital management under the guidance of the HICC provided necessary PPE for the staff as per the exposure risk & working environment and organised training through Online mode on Infection control measures^30^. COVID 19 testing for all In Patients & their bystanders. Staff surveillance team to assess exposure and quarantine of staff and their contacts. In-house testing facility for COVID PCR, Antigen & antibody testing were started. We assessed and facilitated willingness and emotional preparedness of staff in managing the cases well before the first admission ^31^. The audit conducted after all preparations showed a fairly good level of practice. All these might have reduced the chance of exposure to virus during duty or at outside and hence the antibody development in them was low.

## Conclusions

Keeping track of the pattern of development of immunity in the community is part of understanding the illness and forecasting the spread. For the tested HCW, it will boost up morale by ending uncertainty. For the hospital administration it will help in decision making about relative focusing of interventions on patients in general and HCWs. By knowing the immunity status of HCWs, the Institution will be able to contribute authentically to the development of intervention strategies ^32-34^ and guidelines from time to time, besides following the available guidelines. This is a responsibility of a leading academic institution with serving experts, whose expertise will remain dormant otherwise. Being an educational institution, it is obligatory to train all the elements of care delivery to the future generation of health care workers. It includes how to observe, confirm observations, identify the mechanisms behind it and develop intervention strategies with rationale. Understanding the development of personal and herd immunity to a virus infection is hence inevitable. Getting experienced from a small but relevant sample was expected to facilitate larger community study envisaged in peripheral areas Jubilee served.

## Data Availability

Data available on request

